# Data-driven calibration of low-cost wearable motion trackers for gait and dynamic stability measurement

**DOI:** 10.64898/2026.07.13.26357919

**Authors:** Yixuan He, Yuxin Dong, Matthew A. Brodie, Juno Kim, Stephen R. Lord, Yoshiro Okubo

## Abstract

Low-cost inside-out wearable trackers can be deployed at scale to measure body motion, but errors in estimated sensor position propagate through coordinate transformations into derived gait and dynamic-stability metrics. Healthy adults walked on a treadmill at 0.5–2.0 m/s while VIVE Ultimate Tracker (VUT) and Vicon data were recorded. Data-driven calibration models were developed to correct tracker coordinates and to estimate full-body centre of mass (CoM) from a sacrum-only configuration. Agreement with Vicon was assessed using RMSE, mixed-effects Bland-Altman limits of agreement, MAE, and intraclass correlation coefficients. Calibration improved coordinate-level agreement. For gait parameters, model-corrected VUT showed small errors against Vicon (MAE: 0.24–0.71 mm step height, 1.73–4.63 mm step length, 0.15–0.95 mm step width, 0.26–0.88 mm foot clearance). Proxy CoM-derived margin of stability (MoS) agreed excellently with Vicon. For the sacrum-only pipeline, calibration reduced CoM RMSE from 103.65–104.04 mm to 7.55–8.95 mm, and markedly reduced systematic error in stability outcomes, with extrapolated CoM bias decreasing from 172.92 to 0.29 mm and MoS bias from −75.09 to −3.54 mm. Data-driven calibration improved the measurement utility of low-cost VUTs, enabling inexpensive, relatively simple gait and stability measurement from a sacrum-only setup in controlled settings.

## 1. INTRODUCTION

Accurate motion measurement is essential for quantifying gait and dynamic stability in biomechanics, rehabilitation, and related fields [1, 2]. Derived gait and stability metrics, such as step length, step width, step height and margin of stability (MoS), are widely used to quantify human movement [3–6]. Traditionally, high-precision optical marker-based motion capture systems, which rely on infrared cameras and reflective markers, are regarded as “gold standard” for gait analysis due to their spatial accuracy and reliability, often within sub-centimetre ranges (∼1 mm error) [7, 8]. However, these systems are costly, require specialised laboratory environments, and restrict evaluations to controlled indoor settings, limiting their feasibility for continuous or large-scale assessments in real-world contexts [8].

In recent years, wearable and low-cost motion-tracking technologies have emerged as promising alternatives [8]. Devices based on inertial measurement units (IMUs) and inside-out vision-based tracking, originally developed for virtual-reality (VR) body tracking, offer enhanced portability, affordability, and user accessibility [9–11]. Unlike conventional IMUs, inside-out VR trackers provide global six-degree-of-freedom pose estimates, but their accuracy may depend on visual tracking conditions, body attachment, and walking speed [9]. These technologies enable gait monitoring in daily-life environments and hold significant potential for rehabilitation and gait training [9]. Nevertheless, low-cost tracking systems often face challenges such as sensor drift and low spatial accuracy [12, 13]. Consequently, relying on raw tracker measurements for derived gait and stability metrics remains problematic [9, 11]. This gap limits large-scale and longitudinal monitoring outside controlled laboratories.

Data-driven approaches, including machine learning, enable new possibilities for processing sensor data and have shown considerable promise in modelling complex nonlinear dynamics and correcting sensor inaccuracies [14–17]. Specifically, deep learning models, such as convolutional neural networks, Transformer and attention-based models have been successfully applied to time-series data within biomechanics and human activity recognition using wearable sensors [17–21]. By accurately mapping low-cost sensor outputs and target reference data, data-driven calibration models can accurately reconstruct motion trajectories and derive biomechanical parameters [15, 22, 23]. This strategy enables low-cost wearable tracking systems to approximate laboratory-level measurement fidelity while preserving their advantages of low cost and portability [15, 23, 24].

Recent studies have explored the integration of wearable sensors and data-driven approaches for gait estimation, this work has primarily targeted lower-limb kinematic and kinetic variables, such as hip and ankle joint angles and joint moments [25, 26], spatiotemporal gait parameters (e.g., step length and step width), centre of mass (CoM) measures, and discrete gait events (e.g., heel strike and toe off) [23, 24, 27–29]. However, no studies, as yet, have examined dynamic stability parameters such as MoS. Given that gait is inherently dynamic and depends on both the position and velocity of the body’s CoM relative to the base of support (BoS), the extrapolated centre of mass (XCoM) and the associated MoS provide physically grounded metrics for quantifying dynamic balance control during walking [30]. Because this coupling requires velocity estimation and depends on accurate timing, small errors in the foot and sacrum trajectories can be amplified into stability estimates, making valid CoM and MoS estimation from low-cost tracking an accurate clinically relevant test of measurement utility.

Therefore, this study aimed to validate a data-driven calibration approach for low-cost wearable trackers against a Vicon reference system, from coordinate-level error to derived gait and dynamic stability metrics. We further tested whether a sacrum-only setup could estimate full-body CoM sufficiently to derive MoS. We hypothesised that (H1) low-cost wearable trackers would demonstrate acceptable agreement with Vicon for key gait outcomes, and a data-driven calibration would improve agreement for error-sensitive measures; and (H2) that calibration would enable accurate sacrum-only estimation of CoM and CoM-derived MoS across a range of gait speeds. Analyses were structured hierarchically from coordinate error to downstream gait outcomes and CoM-derived stability.

## 2. METHODS

### 2.1. Data collection

This study was conducted at Neuroscience Research Australia (NeuRA) in accordance with the Declaration of Helsinki and approved by the University of New South Wales (UNSW) Human Research Ethics Committee (iRECS6818). Written informed consent was obtained from all participants before participation. Ten participants (men/women: 5/5) were recruited at NeuRA. Participants had a mean age of 32.2 ± 6.5 years (range: 24–44), with an average weight of 66.5 ± 8.0 kg, height of 1.69 ± 0.01 m, and body mass index (BMI) of 23.2 ± 1.8 kg/m² (mean ± standard deviation). Inclusion criteria were: (i) able to communicate in the English language; (ii) aged 20–70 years. Exclusion criteria were: (i) diagnosis of a neurological condition (e.g., Parkinson’s disease, multiple sclerosis, dementia); (ii) inability to walk for 20 minutes without a mobility aid or rest; (iii) history of lower limb, pelvic or vertebral fractures or lower limb joint replacements in the past 6 months; (iv) conditions that prevent exercising (e.g., severe pain, fatigue, exercise intolerance, heel ulcers); (v) advice from a medical practitioner to avoid exercise; (vi) history of dizziness, vertigo and vestibular disorders (e.g., Meniere’s disease).

Participants attended NeuRA for a 1.5-hour laboratory session. This session commenced with assessments of body weight and height. Three trackers were placed on the sacrum and both feet, and a full-body Vicon marker set comprising 39 reflective markers was collected concurrently (Figure 1a and 1b). Tracker-mounted reflective markers enabled physical spatial alignment between systems (Figure 1c and 1d). SteamVR (Steam, Valve, USA) running on a desktop PC served as the platform for collecting VUTs’ data. Participants undertook a 30-second familiarisation walk on a treadmill (M-Gait, Motek®, Netherlands) with a belt speed of 0.5 m/s. Participants then completed four consecutive 1-minute walking tasks at treadmill speeds of 0.5, 1.0, 1.5, and 2.0 m/s. The tasks were performed continuously, with participants notified of each upcoming speed change via a 3-second countdown prompt.

**Figure 1.**
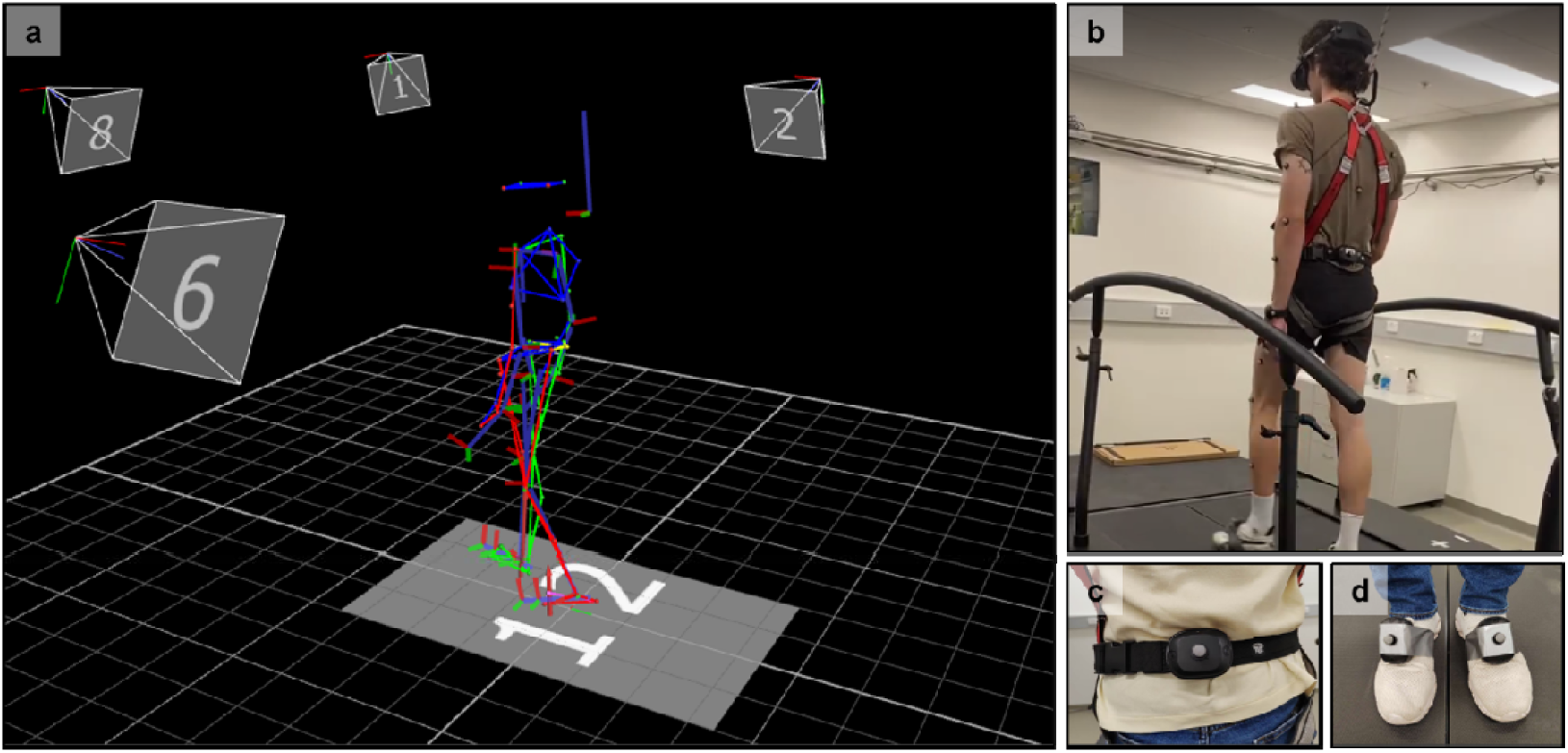
a) full-body markers in the Vicon system, b) a participant walking on the treadmill with a harness, c) Vicon markers and VIVE Ultimate Tracker on the participant’s sacrum, d) Vicon markers and VIVE Ultimate Trackers on the participant’s feet.

An 8-camera Vicon motion capture system (Bonita, Vicon Motion Systems Ltd., Oxford, UK) and Vicon Nexus 2.16.0 x64 software were used to capture three-dimensional (3D) coordinates of the reflective markers at a sampling frequency of 100 Hz. The origin of the Vicon’s coordinate system is the centre of the treadmill surface. An open-source Python script [31] and Visual Studio 2022 (Microsoft, USA) were utilised to collect 3D coordinates from the VUTs during experiments at 100 Hz. The setting and data collection of VUTs were based on the SteamVR platform without additional on-the-fly filtering or smoothing.

Two data-driven calibration model training tasks were performed using synchronised Vicon and VUT trajectories. In Task 1, a data-driven calibration model was trained to correct the raw VUT tracker coordinates to be more consistent with the Vicon marker coordinates at the corresponding tracker locations (sacrum, left foot, and right foot). Coordinate axis directions, spatial magnitude, tracker locations, and treadmill speeds were entered into the model as hand-crafted features. Gait spatiotemporal and dynamic stability parameters were subsequently computed from three trajectory sources (Vicon, raw VUT, and model-corrected VUT) to evaluate whether coordinate calibration improves downstream biomechanical outcomes. Accordingly, the main evaluation in Task 1 focused on gait and proxy-stability outcomes derived from the trajectories, rather than on coordinate correction alone.

In Task 2, the same model architecture was applied to train and estimate the full-body CoM derived from full-body Vicon markers using only the sacrum-mounted VUT trajectory as input [17], and the resulting CoM estimates were used to compute XCoM and MoS and compared with those obtained from raw VUT and the Vicon gold-standard reference. Coordinate axis directions, spatial magnitude, treadmill speeds, and participant pelvis thicknesses were entered into the model as hand-crafted features. Full-body CoM from the Vicon full-body Plug-in-Gait model was computed using a standard segmental approach based on the full-body marker set and anthropometric parameters, yielding a 3D CoM trajectory at 100 Hz as the gold-standard reference in Task 2. For the raw baseline in Tasks 1 and 2, we used the sacrum-mounted VUT position as a proxy for full-body CoM, which represents a common approximation [32, 33], to calculate proxy XCoM and proxy MoS. Task 1 constitutes the primary validation of VUT-derived gait outcomes against Vicon, whereas Task 2 is a secondary analysis designed to test whether CoM-derived stability metrics can be enabled under a minimal sacrum-only configuration.

### 2.2. Data pre-processing

The VUT pose matrix was provided by SteamVR to transform the local coordinate system into the global coordinate system. During synchronisation between the VUT and Vicon coordinate systems, VUT frame loss was identified; to compensate for missing frames, Piecewise Cubic Hermite Interpolating Polynomial (PCHIP) interpolation was applied to the VUT data to match the data length of the Vicon system for each second [34]. The Kabsch algorithm and singular value decomposition [35, 36] were utilised to compute transformations between the SteamVR and Vicon coordinate systems. More detailed procedures are provided in the GitHub repository linked in the Data availability section.

Due to manual operation delays in both Vicon Nexus and the open-source Python script [31] during data collection, a temporal offset emerged between the two datasets. To address this, the first and last 5 seconds were removed from both the Vicon and VUT recordings of each participant at each speed, retaining the central 50 seconds of data. Given the Vicon system’s sampling rate of 100 Hz, this 50-second segment contained 5000 frames. Data were segmented, labelled by participant, speed, location, and direction. All trajectories were expressed in a fixed coordinate system with anterior-posterior (AP), medio-lateral (ML), and vertical (VT) directions. All the scripts for data pre-processing were created in MATLAB R2022a (MathWorks, USA).

### 2.3. Model description

We utilised a data-driven calibration model to reduce spatial errors in low-cost tracker trajectories relative to the Vicon gold-standard reference [16, 19, 20, 23]. In Task 1, the prediction target was the Vicon marker coordinate at the corresponding tracker location. However, in Task 2, the prediction target was the full-body CoM coordinate derived from the Vicon Plug-in-Gait model. The model used a convolutional neural network module with an attention-based Transformer module to map short windows of tri-axial tracker coordinates to the corresponding Vicon-aligned 3D position at the prediction frame (Figure 2) [37]. The convolutional component was used to capture local movement patterns, and the Transformer module was used to model temporal dependencies across the input window. The model thus acts as a calibration stage that maps the raw tracker reading to a corrected estimate of the measurand (3D position in Task 1 and whole-body CoM in Task 2), so that derived biomechanical outcomes, including gait parameters and dynamic stability, remain valid when using low-cost trackers. For each prediction, the model received a fixed-length window of tri-axial tracker coordinates and generated a corrected three-dimensional position for the final frame of that window. Let **x**_*t-L*+1:*t*_ ᗴ ℝ^*L*×3^ denote a fixed-length window of tri-axial tracker coordinates sampled at 100 Hz, where *L* is the sequence length and *t* indexes the final frame. The network learned a mapping *f*_θ_(.) that directly predicted the corrected three-dimensional position at the final frame:

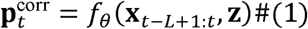

**Figure 2.**
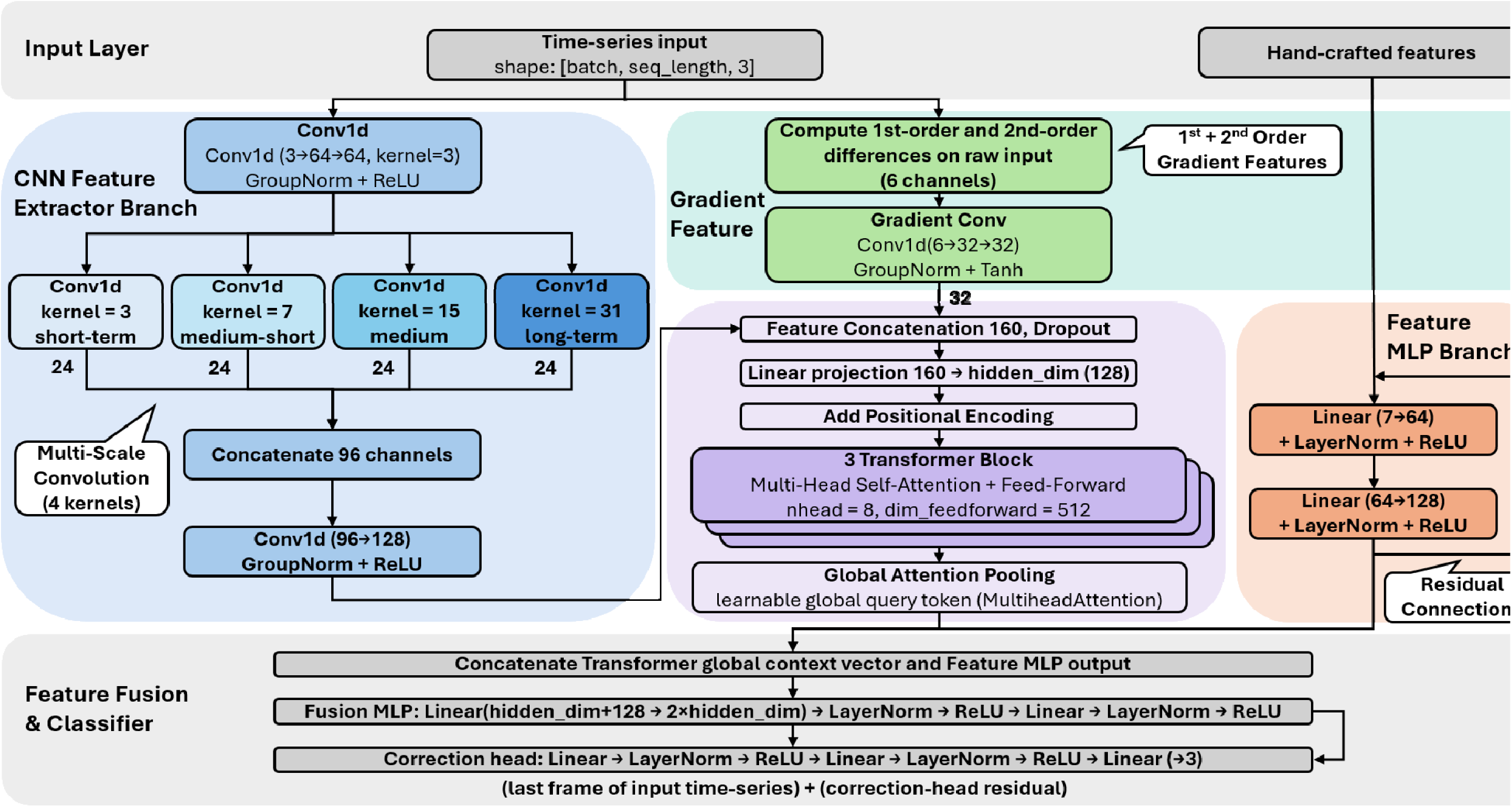
The data-driven calibration model architecture includes a convolutional neural network module and a Transformer module. Coordinate axis directions, spatial magnitude, tracker locations, and speeds were input into the model as hand-crafted features for Task 1. Coordinate axis directions, spatial magnitude, speeds, and participant pelvis thicknesses were input into the model as hand-crafted features for Task 2.

where **z** ᗴ ℝ^*q*^ denotes the auxiliary trial-level features, with *q* depending on the feature set used for each task. Model parameters were optimised by minimising the mean squared error between the corrected coordinates and the Vicon gold-standard reference [19], as shown in equation (2). The primary objective was defined as the mean squared error across all three coordinate dimensions at the prediction frame:

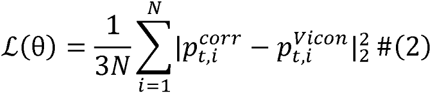

Models were trained to minimise the mean squared error between corrected and gold-standard coordinates at the prediction frame. Leave-one-participant-out cross-validation was used to assess whether the calibration produced consistent biomechanical outcome estimates across participants under the present treadmill protocol (n = 10 for Task 1; n = 8 for Task 2 due to Vicon full-body tracking issues) [38]. Any normalisation or scaling was computed using only the training data within each cross-validation fold and then applied to the held-out test data.

### 2.4. Performance evaluation

Performance was evaluated at the coordinate level and the derived-outcome level. At the coordinate level, accuracy relative to the Vicon gold-standard reference was quantified using three-dimensional root mean squared error (RMSE). RMSE was reported for each of the four treadmill speeds (0.5, 1.0, 1.5, and 2.0 m/s) and each of the three tracker locations (sacrum, left foot, and right foot). RMSE was calculated for two comparisons: (i) the raw VUT coordinates relative to the Vicon gold-standard reference, and (ii) the model-corrected VUT coordinates relative to the Vicon gold-standard reference. This comparison framework allowed direct evaluation of whether the proposed model reduced overall spatial error across all speed × tracker-location combinations (4 speeds × 3 locations). RMSE was defined as:

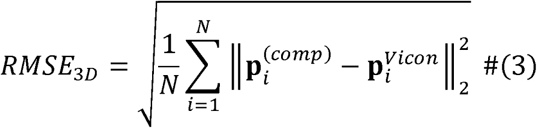

where **p**_*i*_^(Vicon)^ is the Vicon gold-standard coordinate at frame *i*, **p**_*i*_^(comp)^ is the coordinate from the comparison method (either the raw VUT data or the model-corrected VUT data), and *N* is the total number of analysed frames.

At the gait-outcome level, the corrected trajectories were used to derive step height, step length, step width, foot clearance, XCoM, and MoS. Whereas step variables were obtained from the reconstructed foot trajectories, XCoM and MoS were derived from the sacrum and foot trajectories, using sacrum position as a proxy for full-body CoM in accordance with established gait-stability modelling approaches [39]. XCoM, including proxy XCoM, was computed following the inverted pendulum formulation:

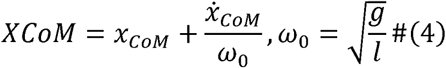

where *x*_CoM_ and *ẋ*_CoM_ are the CoM, or proxy CoM, position and velocity in the direction of interest, *g* is gravitational acceleration, and *l* is the effective pendulum length [30]. The effective pendulum length, *l*, was defined for each participant’s leg length. MoS was defined as the anterior-posterior difference between the BoS boundary and XCoM at heel strike, with positive values indicating that XCoM remained within the BoS boundary. The BoS boundary was estimated from the anterior-posterior direction horizontal distance between the tracker projection and each participant’s heel and toe positions:

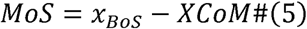

where *x*_*BoS*_ denotes the anterior-posterior position of the relevant BoS boundary at heel strike. These definitions provide a mechanistically grounded index of instantaneous dynamic balance control during walking.

Step height was defined as the maximum vertical distance between the foot tracker and the treadmill surface during swing. Step length and step width were computed from successive foot placements using the AP and ML distances between contralateral foot positions at heel strike, respectively. Heel strikes were detected as local minima in the low-pass-filtered VT foot trajectory using an adaptive, period-constrained peak-finding approach, followed by outlier rejection, and gait cycles were defined as consecutive heel-strike intervals [40]. Subsequently, at least 3444 steps were detected for Task 1, and 2767 steps were detected for Task 2. Foot clearance was quantified as the minimum vertical distance between the foot trajectory and the ground during swing. Dynamic stability was assessed using XCoM and MoS in the AP direction under two calculation modes. XCoM and MoS were computed at heel strike events to provide dynamic stability summaries [30].

The primary agreement was assessed using mixed-effects Bland–Altman limits of agreement (LoA) to account for repeated observations within participants across treadmill speeds [41]; specifically, the between-method differences were modelled with a linear mixed-effects structure (participant as a random effect) to estimate the overall bias and 95% LoA from the relevant variance components. In addition, secondary accuracy and reliability were quantified using mean absolute error (MAE) and the intraclass correlation coefficient (ICC), respectively. ICC was computed using a two-way mixed-effects model for absolute agreement with single measurements. ICC values <0.5 are indicative of poor reliability, 0.5–0.75 indicate moderate reliability, 0.75–0.9 indicate good reliability, and >0.90 indicate excellent reliability [42]. For outcomes, overall ICC and MAE were first computed for the average of each participant’s gait parameters and dynamic stability metrics [42, 43], followed by speed-specific ICC and MAE at 0.5, 1.0, 1.5, and 2.0 m/s. The data-driven calibration model was trained and evaluated on the Katana system, a shared computational cluster at UNSW [44]. Core training settings are summarised in Appendix B, and full source code and hyperparameter files are provided in the GitHub repository listed in Data availability.

## 3. RESULTS

Results are presented as a hierarchical validation of measurement utility: coordinate-level agreement, Task 1 gait and proxy-stability outcomes, and Task 2 sacrum-only CoM-derived stability outcomes.

### 3.1. Coordinate-level accuracy

Across all speeds, calibration improved spatial agreement with Vicon for all tracker locations (Table 1 and Figure 3). Sacrum RMSE decreased from 3.10–3.92 mm (raw VUT) to 1.96–2.67 mm (model-corrected VUT) across speeds. Foot trackers showed larger baseline errors but larger relative gains: right foot RMSE decreased from 7.89–11.58 mm to 4.41–6.82 mm across speeds, and left foot RMSE decreased from 7.63–12.29 mm to 4.42–5.60 mm across speeds. Overall, RMSE decreased from 3.67 to 2.46 mm at the sacrum, 11.00 to 5.91 mm at the right foot, and 10.57 to 5.31 mm at the left foot; across speeds, improvements ranged from 28.39% to 55.25%. Architecture benchmarking is provided in Appendix B.

**Figure 3.**
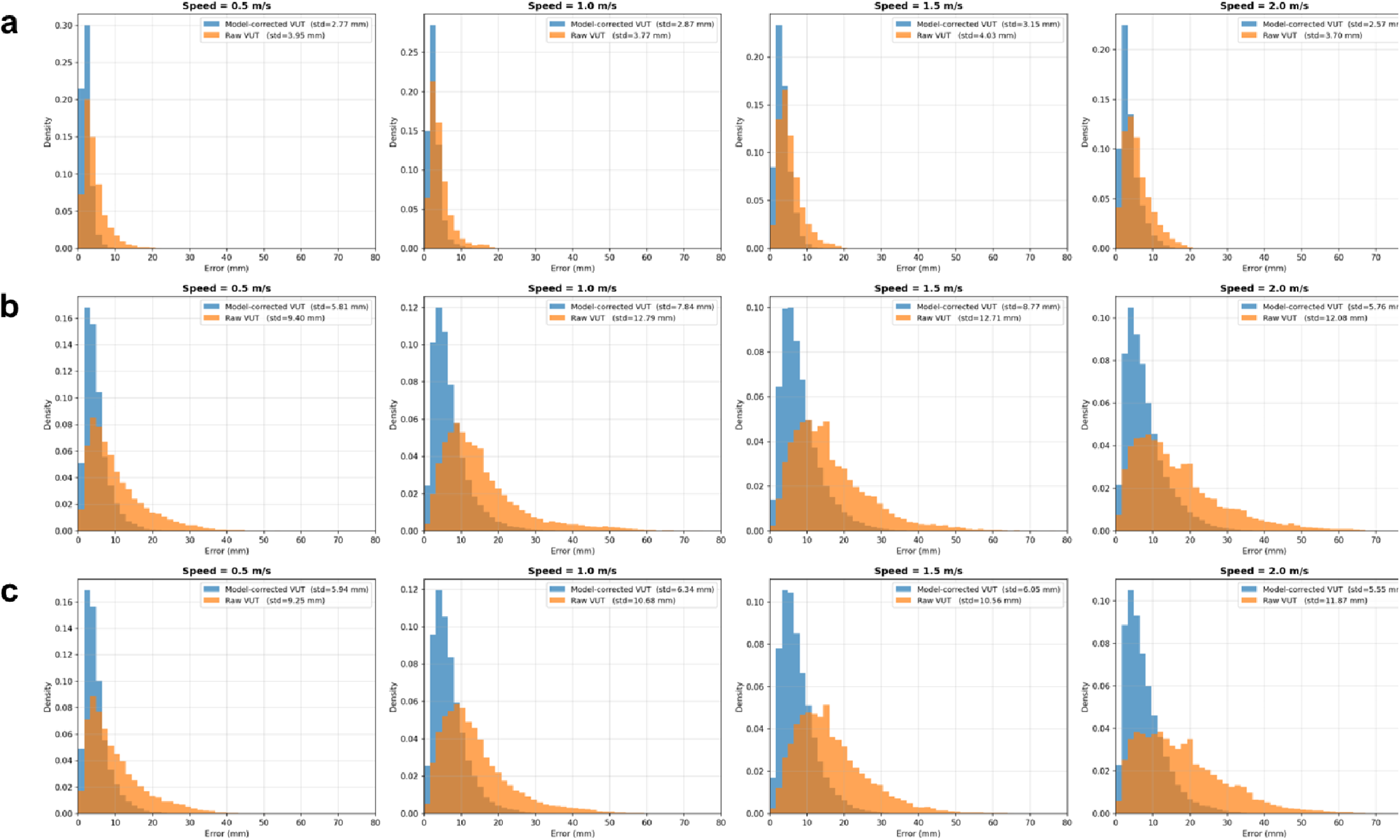
The error distribution of raw VIVE Ultimate Tracker (VUT) versus Vicon, and the model-corrected VUT versus Vicon: a) VUT placed on the sacrum position; b) VUT placed on the right foot position; c) VUT placed on the left foot position.

**Table 1.** Root mean square error (RMSE) of raw VIVE Ultimate Tracker (VUT) and Vicon, model-corrected VUT and Vicon, and the improvement of the two RMSEs.

|  | Treadmill speed | RMSE |  |  |
| --- | --- | --- | --- | --- |
|  |  | Raw VUT and Vicon (mm) | Model-corrected VUT and Vicon (mm) | Improvement (%) |
| Sacrum tracker | Overall | 3.67 | 2.46 | 32.97 |
|  | 0.5 m/s | 3.31 | 1.96 | 40.79 |
|  | 1.0 m/s | 3.10 | 2.22 | 28.39 |
|  | 1.5 m/s | 3.78 | 2.67 | 29.37 |
|  | 2.0 m/s | 3.92 | 2.57 | 34.44 |
| Right foot tracker | Overall | 11.00 | 5.91 | 46.27 |
|  | 0.5 m/s | 7.89 | 4.41 | 44.11 |
|  | 1.0 m/s | 11.13 | 5.98 | 46.27 |
|  | 1.5 m/s | 11.54 | 6.82 | 40.90 |
|  | 2.0 m/s | 11.58 | 5.66 | 51.12 |
| Left foot tracker | Overall | 10.57 | 5.31 | 49.76 |
|  | 0.5 m/s | 7.63 | 4.42 | 42.07 |
|  | 1.0 m/s | 9.82 | 5.39 | 45.11 |
|  | 1.5 m/s | 11.16 | 5.60 | 49.82 |
|  | 2.0 m/s | 12.29 | 5.50 | 55.25 |

### 3.2. Task 1: Gait parameters

For Task 1, the raw VUT preserved excellent agreement with Vicon for step length, step width, foot clearance, and step height, even without the data-driven calibration (ICC: 0.92 to >0.99, Table 2). Across speeds, model-corrected VUT’s MAE was 0.24–0.71 mm for step height, 1.73–4.63 mm for step length, 0.15–0.95 mm for step width, and 0.26–0.88 mm for foot clearance. Mixed-effects Bland-Altman analysis showed small overall bias for the model-corrected VUT gait outcomes, with overall bias of 0.41 mm for step height, -3.67 mm for step length, -0.63 mm for step width, and 0.47 mm for foot clearance (Figure 4 and Appendix A).

**Figure 4.**
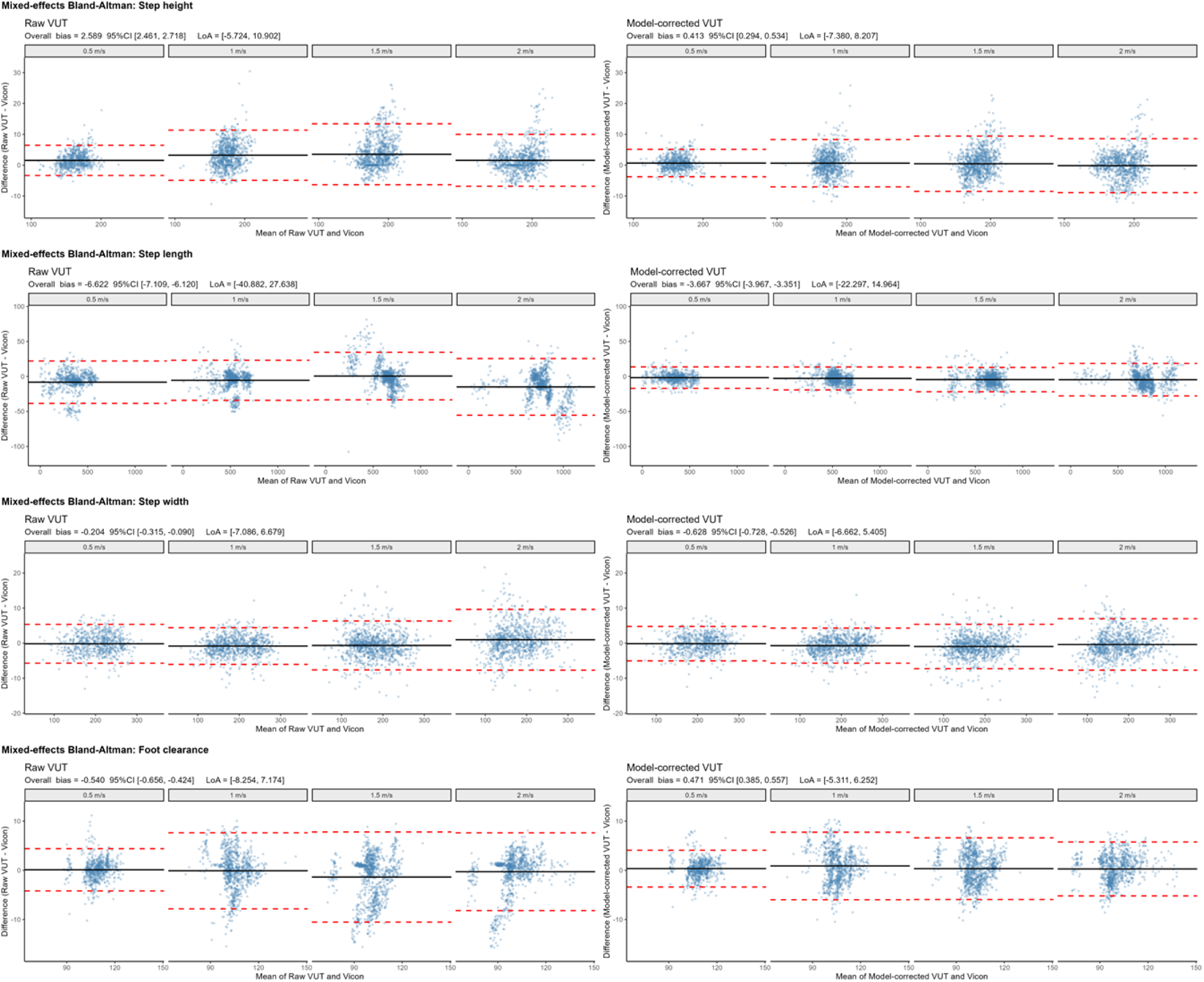
The mixed-effects Bland-Altman plots of gait parameters at different treadmill speeds with millimetre units. CI: confidence interval, LoA: limits of agreement, VUT: VIVE Ultimate Tracker.

**Table 2.** Gait parameters measured by the Vicon, raw VIVE Ultimate Tracker (VUT), and model-corrected VUT at different treadmill speeds.

| Gait parameters | Speed (m/s) | Vicon (mm) | Raw VUT (mm) | Model-corrected VUT (mm) | Vicon vs. raw VUT |  | Vicon vs. Model-corrected VUT |  |
| --- | --- | --- | --- | --- | --- | --- | --- | --- |
|  |  |  |  |  | MAE (mm) | ICC | MAE (mm) | ICC |
| Step height | Overall | 175.94 | 178.46 | 176.38 | 2.52 | 0.97 | 0.44 | 0.99 |
|  | 0.5 | 162.95 | 164.47 | 163.61 | 1.52 | 0.99 | 0.66 | >0.99 |
|  | 1.0 | 174.92 | 178.14 | 175.56 | 3.22 | 0.93 | 0.64 | 0.98 |
|  | 1.5 | 183.35 | 187.15 | 184.06 | 3.80 | 0.94 | 0.71 | 0.98 |
|  | 2.0 | 182.54 | 184.06 | 182.30 | 1.52 | 0.99 | 0.24 | 0.99 |
| Step length | Overall | 574.59 | 567.57 | 571.17 | 7.02 | >0.99 | 3.42 | >0.99 |
|  | 0.5 | 351.92 | 343.74 | 350.19 | 8.18 | 0.97 | 1.73 | >0.99 |
|  | 1.0 | 531.25 | 525.76 | 528.40 | 5.49 | 0.97 | 2.85 | >0.99 |
|  | 1.5 | 630.34 | 630.88 | 625.86 | 0.54 | 0.99 | 4.48 | >0.99 |
|  | 2.0 | 784.86 | 769.91 | 780.23 | 14.95 | 0.99 | 4.63 | >0.99 |
| Step width | Overall | 187.41 | 187.23 | 186.86 | 0.18 | >0.99 | 0.55 | >0.99 |
|  | 0.5 | 203.99 | 203.80 | 203.84 | 0.19 | >0.99 | 0.15 | >0.99 |
|  | 1.0 | 191.94 | 191.11 | 191.23 | 0.83 | >0.99 | 0.71 | >0.99 |
|  | 1.5 | 179.27 | 178.61 | 178.32 | 0.66 | >0.99 | 0.95 | >0.99 |
|  | 2.0 | 174.44 | 175.40 | 174.06 | 0.96 | >0.99 | 0.38 | >0.99 |
| Foot clearance | Overall | 103.87 | 103.43 | 104.33 | 0.44 | 0.95 | 0.46 | 0.97 |
|  | 0.5 | 108.93 | 109.03 | 109.28 | 0.10 | >0.99 | 0.35 | 0.99 |
|  | 1.0 | 105.00 | 104.91 | 105.88 | 0.09 | 0.93 | 0.88 | 0.93 |
|  | 1.5 | 101.42 | 99.97 | 101.75 | 1.45 | 0.92 | 0.33 | 0.97 |
|  | 2.0 | 100.13 | 99.82 | 100.39 | 0.31 | 0.95 | 0.26 | 0.98 |
| MAE: | mean | absolute | error, | ICC: | intraclass | correlation | coefficient. |  |

### 3.3. Task 1: Dynamic stability using proxy CoM

For proxy CoM stability outcomes, agreement with Vicon was already high for proxy XCoM, whereas data-driven calibration provided clearer improvement for proxy MoS, mainly through reduced MAE and bias (Table 3, Figure 5, and Appendix A). Model-corrected VUT’s proxy XCoM showed an overall bias of -3.45 mm with LoAs of -28.46 to 21.55 mm, which was similar to the raw VUT proxy XCoM (Figure 5 and Appendix A). For proxy MoS, the model-corrected VUT reduced the absolute overall bias from 3.54 mm to 1.20 mm and slightly narrowed the overall LoA width, mainly through an improved lower LoA from -31.31 to -24.34 mm.

**Figure 5.**
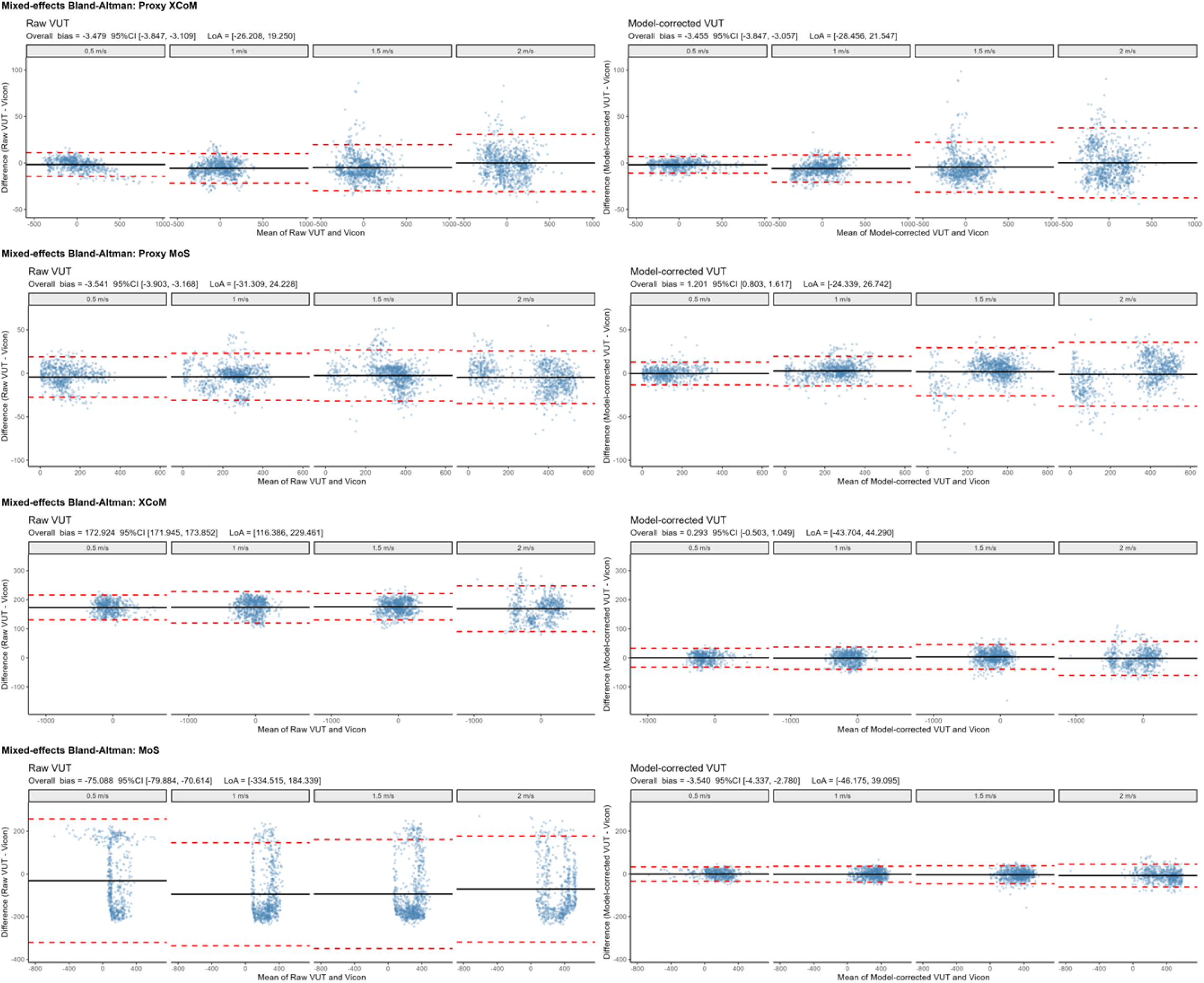
The mixed-effects Bland-Altman plots of dynamic stability metrics at different treadmill speeds with millimetre units. CI: confidence interval, LoA: limits of agreement, VUT: VIVE Ultimate Tracker.

**Table 3.** Dynamic stability measured by the Vicon, raw VIVE Ultimate Tracker (VUT), and model-corrected VUT at different treadmill speeds.

| Parameters | Speed (m/s) | Vicon (mm) | Raw VUT (mm) | Model-corrected VUT (mm) | Vicon vs. raw VUT |  | Vicon vs. Model-corrected VUT |  |
| --- | --- | --- | --- | --- | --- | --- | --- | --- |
|  |  |  |  |  | MAE (mm) | ICC | MAE (mm) | ICC |
| Proxy XCoM | Overall | -7.94 | -11.03 | -10.96 | 3.09 | >0.99 | 3.02 | >0.99 |
|  | 0.5 | 6.99 | 5.38 | 5.15 | 1.61 | >0.99 | 1.84 | >0.99 |
|  | 1.0 | -19.56 | -25.32 | -25.53 | 5.76 | >0.99 | 5.97 | >0.99 |
|  | 1.5 | -17.11 | -22.12 | -21.58 | 5.01 | >0.99 | 4.47 | >0.99 |
|  | 2.0 | -2.07 | -2.05 | -1.88 | 0.02 | >0.99 | 0.19 | >0.99 |
| Proxy MoS | Overall | 256.02 | 252.26 | 256.85 | 3.76 | >0.99 | 0.83 | >0.99 |
|  | 0.5 | 136.19 | 132.03 | 136.05 | 4.16 | 0.97 | 0.14 | >0.99 |
|  | 1.0 | 250.74 | 246.81 | 253.37 | 3.93 | 0.99 | 2.63 | >0.99 |
|  | 1.5 | 323.22 | 320.76 | 325.08 | 2.46 | 0.99 | 1.86 | >0.99 |
|  | 2.0 | 313.92 | 309.45 | 312.9 | 4.47 | >0.99 | 1.02 | >0.99 |
| XCoM | Overall | -113.94 | 59.30 | -113.55 | 173.24 | 0.55 | 0.39 | >0.99 |
|  | 0.5 | -131.75 | 41.48 | -131.62 | 173.23 | 0.19 | 0.13 | >0.99 |
|  | 1.0 | -119.01 | 54.98 | -119.93 | 173.99 | 0.43 | 0.92 | >0.99 |
|  | 1.5 | -101.74 | 74.01 | -98.42 | 175.75 | 0.48 | 3.32 | >0.99 |
|  | 2.0 | -101.74 | 67.8 | -102.9 | 169.54 | 0.80 | 1.16 | >0.99 |
| MoS | Overall | 283.67 | 210.57 | 280.59 | 73.10 | 0.65 | 3.08 | >0.99 |
|  | 0.5 | 159.55 | 128.36 | 159.25 | 31.19 | 0.09 | 0.30 | >0.99 |
|  | 1.0 | 301.91 | 207.04 | 300.51 | 94.87 | 0.28 | 1.40 | >0.99 |
|  | 1.5 | 331.76 | 237.78 | 328.01 | 93.98 | 0.30 | 3.75 | >0.99 |
|  | 2.0 | 349.71 | 277.47 | 342.31 | 72.24 | 0.79 | 7.40 | >0.99 |

XCoM: extrapolated centre of mass, MoS: margin of stability, MAE: mean absolute error. Proxy XCoM/MoS: Vicon/VUT sacrum position as CoM approximation (Task 1), XCoM/MoS: Vicon CoM is from full-body Plug-in-Gait model, raw VUT CoM was approximated from the sacrum position, whereas model-corrected CoM was estimated from the sacrum-mounted VUT trajectory (Task 2).

### 3.4. Task 2: CoM estimation for dynamic stability

In Task 2, sacrum-only calibration reduced CoM RMSE from 103.65–104.04 mm to 7.55–8.95 mm across speeds, with the overall RMSE decreasing from 103.86 mm to 8.24 mm (Table 4 and Figure 6). This improvement yielded near-Vicon XCoM and MoS estimates (ICC: >0.99, Table 3, Figure 5, and Appendix A). Mixed-effects Bland-Altman analysis showed that model-corrected VUT markedly reduced the systematic error observed with raw VUT stability outcomes: XCoM bias decreased from 172.92 mm to 0.29 mm, and MoS bias decreased from -75.09 mm to -3.54 mm. The corresponding LoA narrowed from 116.39 to 229.46 mm to -43.70 to 44.29 mm for XCoM, and from -334.52 to 184.34 mm to -46.17 to 39.09 mm for MoS.

**Figure 6.**
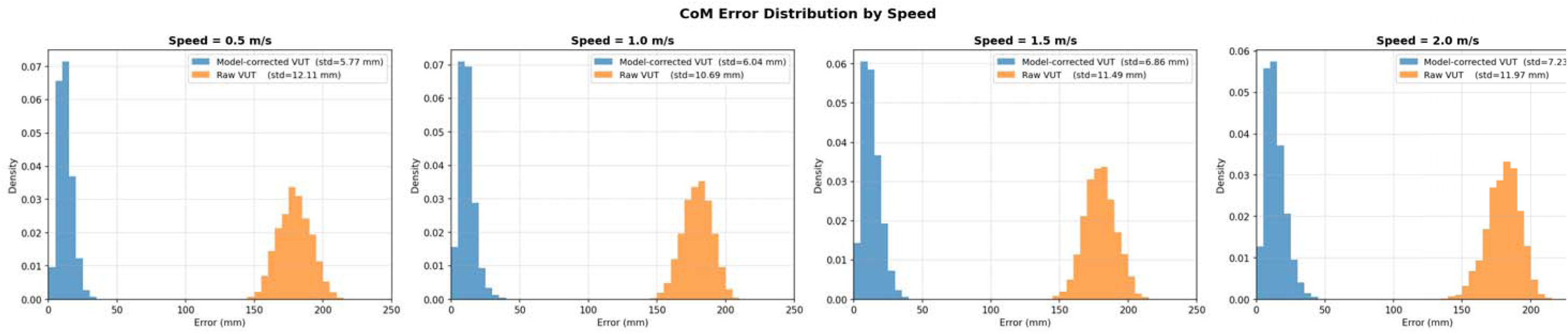
Centre of mass error distribution of raw VIVE Ultimate Tracker (VUT) versus Vicon, and the model-corrected VUT versus Vicon by speed.

**Table 4.** Root mean square error (RMSE) of the centre of mass of raw VIVE Ultimate Tracker (VUT) and Vicon, the centre of mass of the model-corrected VUT and Vicon, and the improvement of the two RMSEs.

| Treadmill speed | RMSE |  |  |
| --- | --- | --- | --- |
|  | Raw VUT and Vicon (mm) | Model-corrected VUT and Vicon (mm) | Improvement (%) |
| Overall | 103.86 | 8.24 | 92.07 |
| 0.5 m/s | 103.65 | 7.78 | 92.49 |
| 1.0 m/s | 103.65 | 7.55 | 92.72 |
| 1.5 m/s | 103.73 | 8.46 | 91.84 |
| 2.0 m/s | 104.04 | 8.95 | 91.40 |

## 4. DISCUSSION

This study aimed to validate a data-driven calibration approach for low-cost inside-out wearable motion tracking against a Vicon gold-standard reference. To this end, we evaluated whether calibration reduces coordinate error and preserves agreement for error-sensitive derived outcomes, including CoM-derived XCoM and MoS. Two principal findings emerged. First, the data-driven calibration consistently reduced coordinate-level error across tracker locations and different walking speeds, with the largest relative gains for the foot trackers (40.90–55.25%) and robust improvements for the sacrum tracker (RMSE reduction: 28.39–40.79%). Second, when applied to sacrum-only input to estimate whole-body CoM, the data-driven calibration reduced CoM RMSE from 103.65–104.04 mm to 7.55–8.95 mm and substantially corrected the systematic bias in CoM-derived stability outcomes. In particular, XCoM bias decreased from 172.92 mm to 0.29 mm, and MoS bias decreased from -75.09 mm to -3.54 mm.

Together, these results suggest that data-driven calibration can translate low-cost wearable tracking into valid derived gait and stability measurements with minimal setup. These high ICCs are similar to those reported in previous studies [9], but should be interpreted alongside MAE and Bland–Altman LoAs, because ICC is sensitive to the variance structure of the data. In fixed-speed treadmill walking, large between-condition (speed) variance and relatively low stride-to-stride variability can yield high ICCs even when small systematic biases remain [42].

### 4.1. Trajectory enhancement and gait parameters

The greatest coordinate-level improvements were observed in the foot trackers, which also presented the largest baseline error. Foot trackers undergo larger accelerations, rapid direction reversals near heel strike and toe off, and frequent changes in visibility when tracked using the inside-out camera vision-based system. Even small systematic distortions in local foot motion are amplified when derived outcomes depend on precise extrema (e.g., peak swing height) or when numerical differentiation is applied [27]. This supports calibration when low-cost trackers are used for derived outcomes sensitive to local extrema and differentiation [14, 24]. Such speed- and motion-dependent degradation has also been reported in prior comparisons of VIVE trackers against optical motion capture, particularly under dynamic movements and varying tracking configurations [9, 11, 45].

Coordinate improvements translated most clearly into step height. Model-corrected VUT’s step height error remained below 1 mm across speeds, with excellent ICCs. Bland-Altman bias also decreased from 2.59 mm with raw VUT to 0.41 mm after modelling. Because step height is a vertical swing-phase measure, it is especially vulnerable to drift, local distortion, and attachment-related error in low-cost tracking systems; correcting these vertical errors is therefore why step height showed the clearest improvement.

For step length, the data-driven calibration reduced overall bias from -6.62 mm to -3.67 mm and narrowed the overall LoAs from -40.88 to 27.64 mm to -22.30 to 14.96 mm. However, the improvement was less uniform across speeds than for step height. An interpretation is that step length is an event-dependent measure: it depends on the timing and identification of gait events, so residual phase shifts or small contact-phase errors can degrade agreement even when overall trajectory RMSE improves.

### 4.2. CoM estimation and dynamic stability

The proxy-CoM analysis and the sacrum-only CoM estimation analysis addressed different levels of measurement difficulty: the former assessed whether sacrum position can support simplified stability estimates, whereas the latter assessed whether a single tracker could recover full-body CoM-derived stability metrics. The proxy CoM-derived XCoM remained highly consistent with Vicon even without data-driven calibration, whereas proxy MoS showed a clearer benefit from modelling. Bland-Altman results indicated that proxy XCoM bias was almost unchanged after modelling, but proxy MoS bias shifted closer to zero, and the lower limit of agreement improved. This suggests that sacrum-position proxy measures may be sufficient for some XCoM estimates under fixed-speed treadmill walking, consistent with previous studies [32, 39], whereas MoS is more sensitive to small position and velocity errors. This is plausible because XCoM and MoS depend on CoM velocity [30], and MoS is evaluated at heel-strike instants relative to the BoS boundary; therefore, small signal-processing or event-detection errors can amplify variability [29, 46]. Future work could improve robustness by incorporating event-aware estimation (e.g., joint modelling of gait events and trajectory calibration) [47].

The sacrum-only pipeline produced the clearest measurement benefit. Raw VUT-derived CoM differed substantially from Vicon, and this propagated into large systematic errors in XCoM and MoS. After modelling, CoM RMSE decreased to 7.55–8.95 mm, XCoM bias was reduced to near zero, and MoS LoAs narrowed markedly. For Task 2, the raw VUT-derived XCoM error was dominated by a large physical offset, whereas MoS showed both large bias and wide LoAs; calibration therefore improved both the centring and interpretability of CoM-derived stability estimates. This large improvement is expected because a single sacrum-mounted tracker does not directly measure whole-body segmental CoM and is additionally affected by global-frame drift and pelvis-to-whole-body mapping differences; the data-driven calibration model effectively learns this mapping under the present fixed-speed treadmill walking conditions [17]. Previous studies have shown that a single sacrum-mounted sensor can be related to CoM displacement during treadmill walking, supporting the plausibility of low-back signals as CoM proxies when carefully processed [28, 32, 39]. In biomechanics, XCoM and MoS are mechanistically grounded metrics that incorporate both position and velocity, and their clinical relevance stems from linking full-body dynamics to balance control [2, 30]. However, accurate CoM estimation typically requires full-body markers or multi-sensor fusion [28].

Our results indicate that raw sacrum-mounted VUT data alone did not achieve good agreement with the gold-standard reference for full-body CoM-derived dynamic stability (XCoM ICC: 0.19–0.80; MoS ICC: 0.09–0.79), even if sacrum position can be a proxy for CoM [28, 32, 39], but when combined with data-driven calibration [17], it can approximate Vicon-level XCoM and MoS (ICC: >0.99). Data-driven calibration approaches that map wearable signals to CoM-derived kinematics further support this bridge to the biomechanics paradigm [17, 48]. This substantially reduces setup burden and may improve scalability for clinical screening, home-based monitoring, and longitudinal tracking in populations at risk of falls [8, 24]. Reducing CoM state error is crucial for obtaining usable MoS, because MoS integrates both position and velocity information at contact transitions [2, 30]. The observed near-gold-standard agreement suggests that the calibration effectively controls this amplification. This finding addresses a central bottleneck in real-world stability biomechanics: CoM-derived metrics are informative but historically difficult to measure outside the lab [2, 24]. Enabling them from a single low-cost tracker meaningfully lowers the barrier to mechanistic balance assessment at scale.

### 4.3. Limitations and future work

Several limitations should be considered. First, the participant sample size was modest (n = 10 for Task 1 and n = 8 for Task 2 due to Vicon full-body tracking issues), internal participant-wise evaluation within a modest cohort may have led to mildly optimistic estimates, and the model appears to have limited transferability to an unseen participant [15]. Second, participants were healthy adults walking on a treadmill at fixed speeds; treadmill gait differs from overground gait in variability, speed regulation, and foot-placement strategies, so transfer to real-world environments remains to be tested [9]. Third, the VT direction of CoM did not perform well with the data-driven calibration model. Since the calculation of dynamic stability in this study only considers the horizontal projection, VT-direction errors did not directly enter the XCoM and MoS calculations in this study; thus, the problem of poor accuracy in the VT direction was not optimised [30, 48]. Fourth, Appendix B benchmarks CNN, Transformer, and CNN–Transformer architectures, but broader comparisons with simpler models such as linear regression or LSTM remain for future work [15]. Finally, tracker placement, attachment stability, SteamVR tracking conditions, and between-session recalibration were not systematically tested; therefore, repeatability across days, operators, and environments remains unknown [11]. Moreover, all recordings used a single fixed laboratory scene; because inside-out tracking relies on environmental visual features, scene articulation may affect tracking quality, particularly in the vertical direction relevant to step height, and was not evaluated here. The extent to which this calibration can be trusted as a measurement tool beyond the present treadmill protocol therefore remains to be established.

Future studies should validate performance in larger, more diverse cohorts, including older people and clinical populations, and evaluate overground walking, turning, and dual-task conditions where dynamic stability is challenged. They should also assess axis-specific errors and integrate event-aware learning, such as multi-task prediction of heel strike and toe off alongside trajectory calibration.

## 5. CONCLUSION

This study demonstrated that data-driven calibration improves the measurement utility of low-cost VIVE Ultimate Trackers under treadmill walking. Calibration reduced coordinate-level error across tracker locations and produced accurate derived gait parameters, while the largest benefit was observed in the sacrum-only CoM pipeline, where CoM RMSE decreased from approximately 104 mm to below 9 mm and large systematic errors in XCoM and MoS were corrected. This framework is not intended to replace optical motion capture, but may provide a lower-burden, reference-traceable measurement pathway for gait and dynamic-stability metrics when laboratory-grade systems are unavailable.

## Data Availability

De‑identifiable participant data (from individuals who consented to data sharing) and the MATLAB scripts used in this study are available in the following public GitHub repository: https://github.com/Yixuan-NeuRA/Validation-of-VIVE-and-VICON. The data-driven calibration model Python scripts used in this study are available in the following public GitHub repository: https://github.com/Yixuan-NeuRA/VIVIAN-VIVE-and-VICON-Integration-for-Adaptive-Network. Hyperparameter configuration files corresponding to Appendix B are included in the model repository.

https://github.com/Yixuan-NeuRA/VIVIAN-VIVE-and-VICON-Integration-for-Adaptive-Network

## ACKNOWLEDGEMENTS

We would like to thank Mr George Mitri for his contribution during the technical development of the VR system and Ms Carly Chaplin for her contribution to data collection and processing. We thank all study participants who volunteered to participate in this study. This research includes computations using the computational cluster Katana supported by Research Technology Services at UNSW Sydney.

## CONTRIBUTORSHIP

Y.H. – Conceptualisation, software, methodology, formal analysis, data curation, visualisation, writing – original draft.

Y.D. – Data curation, visualisation, writing – review & editing.

M.A.B. – Conceptualisation, supervision, writing – review & editing.

J.K. – Conceptualisation, software, supervision, writing – review & editing.

S.R.L. – Conceptualisation, supervision, writing – review & editing.

Y.O. – Conceptualisation, methodology, validation, investigation, resources, supervision, project administration, funding acquisition, writing – review & editing.

## Ethics

Ethical approval was obtained from the Human Research Ethics Committee of the University of New South Wales on 21/10/2024, with Reference Number iRECS6818. Participants provided written consent.

## Declaration of AI use

We used AI in the writing process to improve the readability and language of our manuscript.

## Data accessibility

De-identifiable participant data (from individuals who consented to data sharing) and the MATLAB scripts used in this study are available in the following public GitHub repository: https://github.com/Yixuan-NeuRA/Validation-of-VIVE-and-VICON. The data-driven calibration model Python scripts used in this study are available in the following public GitHub repository: https://github.com/Yixuan-NeuRA/VIVIAN-VIVE-and-VICON-Integration-for-Adaptive-Network. Hyperparameter configuration files corresponding to Appendix B are included in the model repository.

